# Pulmonary ^129^Xe Magnetic Resonance Gas-exchange Abnormalities in Moderate-Severe Asthma

**DOI:** 10.1101/2024.08.01.24311378

**Authors:** Sam Tcherner, Ali Mozaffaripour, Alexander M Matheson, Alexander Biancaniello, Cory Yamashita, Grace Parraga

**Affiliations:** Robarts Research Institute, Western University, London, Canada; Department of Medical Biophysics, Western University, London, Canada; School of Biomedical Engineering, Western University, London, Canada; Division of Respirology, Department of Medicine; Western University, London, Canada

## Abstract

**BACKGROUND:** Asthma is recognized as an inflammatory disease of the airways, but inflammation may also affect the parenchyma and pulmonary vasculature. Hyperpolarized ^129^Xe MRI and MR spectroscopy (MRS) provide a way to quantify the transfer of gas from the airways through the alveolar membrane and its binding to hemoglobin in the red blood cells (RBC) of the pulmonary microvasculature. The vast majority of ^129^Xe MRS studies have investigated interstitial lung disease and the ratio of ^129^Xe binding to the RBC and ^129^Xe present in the alveolar membrane, (RBC:membrane) which is a surrogate of oxygen gas-transfer to the blood. We wondered if ^129^Xe RBC:membrane would differ in asthma patients as compared to healthy volunteers because of recent work showing abnormally diminished pulmonary vascular small-vessel structure in severe-asthma.

**RESEARCH QUESTION:** Do ^129^Xe MRI gas-transfer measurements differ significantly in patients with moderate-severe asthma?

**STUDY DESIGN AND METHODS:** In this retrospective study, healthy (NCT02484885) and asthma (NCT04651777; NCT02351141) participants were evaluated who provided written informed consent.

**RESULTS:** Thirty-one participants with asthma (mean age=55 years ±18; 22 females) and 32 healthy volunteers (mean age=31 years ±14; 12 females) with ^129^Xe MRS were evaluated. FEV_1_, VDP and DL_CO_/K_CO_ were significantly different in asthma compared to healthy participants. Age-corrected ^1^RBC:membrane was significantly different in moderate-severe asthma (0.32±0.09) as compared to healthy participants (0.47±0.12, *P*=.01). RBC:membrane was significantly related to pulse-oximetry hemoglobin estimates (ρ=.29; *P*=.04) and DL_CO_ (ρ=.71; *P*<.001). Significant relationships between ^129^Xe RBC:membrane and age were observed in healthy (ρ=-.55; *P*=.002) and asthma participants (ρ=-0.49; *P*=.006), adjusted for sex. A significant ANCOVA model also revealed the influence of age (*P*=.002), sex (*P*<.001), hemoglobin (*P*=.003) and asthma status (*P*=.02) on RBC:membrane.

**INTERPRETATION:** ^129^Xe RBC:membrane values were significantly different in moderate-severe asthma compared to healthy volunteers and were explained by age, sex, hemoglobin, and asthma status.

## INTRODUCTION

Although asthma is recognized as an inflammatory airway disease, other lung compartments such as the parenchyma and pulmonary vasculature may be modified directly or indirectly, by inflammation.^1-3^ For example, evidence from cross-sectional and longitudinal studies in severe asthma suggested pulmonary vascular differences in the small vessel volume fraction.^4,5^ Reversal of these abnormalities was reported in asthma patients following biologic therapy, as evidenced by a redistribution of blood volume from larger to smaller vessels.^5^ It is possible that some of these abnormalities stem from inflammation and pulmonary vascular remodeling.^2,3^

Hyperpolarized ^129^Xe MRI and MR spectroscopy (MRS) provide a way to quantify the transfer of gas from the airways through the alveolar membrane and its binding to hemoglobin in the red blood cells (RBC) of the pulmonary microvasculature. In these studies, the ratio of ^129^Xe binding to the RBC to ^129^Xe which has participated in transmembrane diffusion into the alveolar membrane, (RBC:membrane) is utilized as a surrogate of oxygen gas-transfer to the blood.^6^ Recent work revealed the reproducibility and age-dependence of RBC:membrane values.^7-10^ The vast majority of such ^129^Xe “multi-compartment” studies have investigated interstitial lung disease,^11-15^ mainly because RBC:membrane is a sensitive marker of alveolar membrane thickening and fibrosis.^16^ In a small number of patients with asthma, a pilot study revealed both abnormally increased and abnormally decreased RBC:membrane values, without RBC:membrane post-bronchodilator response.^17^

Because of these previous contradictory findings, our objective was to acquire and evaluate ^129^Xe MRS gas-exchange measurements in patients with moderate-severe asthma and compare these directly with healthy volunteers. We also aimed to measure the quantitative relationships of RBC:membrane measurements with age, sex, asthma severity, diffusing-capacity of the lung for carbon-monoxide and hemoglobin values.

## METHODS

### Study participants and Design

We retrospectively evaluated 31 participants with asthma and 32 healthy volunteers who provided written informed consent (NCT02484885; NCT04651777; NCT02351141) to pulmonary function tests and MRI. Inclusion criteria for participants with asthma consisted of male and female non-smokers, 18-75 years of age and documented diagnosis of asthma and treated with low-to-high dose ICS/LABA. Healthy volunteers were males and females 18-85 years of age, with ≤1 pack year smoking history and no previous diagnosis or history of chronic respiratory disease.

### Pulmonary Function Tests

Spirometry and DL_CO_ measurements were undertaken according to American Thoracic Society guidelines using a MedGraphic Elite Series system (MedGraphics; St. Paul, MN). Participants with asthma withheld short-acting β-agonists for 6 hours, and long-acting β-agonists for 12 hours prior to study visits,^18^ and completed the Asthma Control Questionnaire (ACQ-6),^19^ Asthma Quality-of-Life Questionnaire (AQLQ),^20^ and St. George’s Respiratory Questionnaire (SGRQ).^21^

### MRI Acquisition and Analysis

^129^Xe gas was polarized to 30% to 55% (XeniSpin 9820; Polarean, Durham, NC, USA). Anatomic (^1^H) MRI, functional (^129^Xe) MRI and MRS were acquired at 3.0 Tesla (Discovery MR750; GE Healthcare, Milwaukee, WI, USA), as previously described.^22^ Spectroscopic measurements were reported as normalized ratios (RBC:membrane, Membrane:gas, RBC:gas) using the area-under-the-curve values from the corresponding RBC, membrane and gas resonance peaks.

### Statistical Analysis

SPSS Statistics version 29.0 (IBM) was used for analysis. Normality was assessed with Shapiro-Wilk tests, and non-parametric tests were used for non-normal data. Differences between participant groups were assessed with independent samples t-tests or Mann-Whitney U-tests. Analysis of covariance (ANCOVA) was used to generate significant models explaining RBC:membrane values. Results were considered statistically significant if the probability of a Type I error was less than 5% (*P*<.05).

## RESULTS

Table 1 shows demographic, pulmonary function, and imaging measurements for 31 participants with asthma (mean age, 56 years ±17; 21 females) and 32 healthy participants (mean age, 31 years ±14; 12 females). The two groups were significantly different for age (*P*<.001), BMI (*P*=.009), FEV_1_ (*P*=.001), FEV_1_/FVC (*P*=.004), DL_CO_ (*P*=.01), K_CO_ (*P*=.003) and VDP (*P*<.001). Mean ^129^Xe RBC:membrane was significantly different when age-adjusted in moderate-severe asthma (0.32±0.09) as compared to the healthy volunteers (0.47±0.12; *P*=.01).

**Table 1.** Demographic and Clinical Characteristics.

| Parameter Mean (SD) | Healthy (n=32) | Asthma (n=31) | P |
| --- | --- | --- | --- |
| Age years | 31 (14) | 56 (17) | **<.001** |
| BMI kg/m<sup>2</sup> | 25 (4) | 29 (7) | **.009** |
| Female n (%) | 12 (38) | 21 (68) |  |
| GINA-4 n (%) | – | 25 (81) |  |
| GINA-5 n (%) | – | 6 (19) |  |
| Pack-years |  |  |  |
| Mean | 0 (0) | 7 (14) | **<.001** |
| Median [IQR] | 0 [0–0] | 0 [0–7] |  |
| Duration of asthma years |  |  |  |
| Mean | 0 (0) | 28 (20) | **<.001** |
| Median [IQR] | 0 [0–0] | 26 [10–39] |  |
| SpHb g/dL | 15.3 (1.4) | 14.8 (1.6) | .2 |
| SpO<sub>2</sub> % | 98 (1) | 97 (2) | **.01** |
| FEV<sub>1</sub> %<sub>pred</sub> | 92 (13) | 79 (17) | **.001** |
| FVC %<sub>pred</sub> | 97 (14) | 91 (12) | .09 |
| FEV<sub>1</sub>/FVC %<sub>pred</sub> | 95 (8) | 85 (13) | **.004** |
| DL<sub>CO</sub> mL/min/mmHg | 34 (9) | 23 (9) | **.003** |
| DL<sub>CO</sub> %<sub>pred</sub> | 121 (19) | 96 (28) | **.01** |
| VA (L) | 5.8 (1.4) | 5.1 (1.6) | .2 |
| VA %<sub>pred</sub> | 103 (14) | 96 (18) | .3 |
| K<sub>CO</sub> mL/min/mmHg/L | 5.8 (0.9) | 4.6 (1.3) | **.004** |
| K<sub>CO</sub> %<sub>pred</sub> | 119 (18) | 99 (17) | **.003** |
| ACQ-6 | – | 1.5 (1.0) |  |
| AQLQ | – | 5.3 (1.0) |  |
| SGRQ | – | 37 (21) |  |
| VDP % | 1 (1) | 8 (8) | **<.001** |
| <sup>129</sup>Xe RBC:Membrane | 0.47 (0.12) | 0.32 (0.09) | **.01\*** |
| <sup>129</sup>Xe Membrane:Gas | 0.99 (0.26) | 0.89 (0.36) | .5* |
| <sup>129</sup>Xe RBC:Gas | 0.47 (0.19) | 0.30 (0.16) | .08* |

n=11 for Asthma DL<sub>CO</sub>, VA, K<sub>CO</sub>; n=12 for Healthy VDP; n=23 for Asthma SpHb; n=26 for Healthy SpHb; n=27 for Asthma SGRQ; n=28 for Healthy DL<sub>CO</sub>, VA, K<sub>CO</sub>.
P=Significance values for independent samples t-test; P\*=Significance values for ANCOVA adjusted for age. BMI=body mass index; GINA=Global Initiative for Asthma; IQR=interquartile range; SpHb=continuous total hemoglobin; SpO<sub>2</sub>=peripheral capillary oxygen saturation; FEV<sub>1</sub>=forced expiratory volume in 1 second; %<sub>pred</sub>=percent of predicted value; FVC=forced vital capacity; DL<sub>CO</sub>=diffusing capacity of the lungs for carbon monoxide; VA=alveolar volume; K<sub>CO</sub>=transfer coefficient of the lung for carbon monoxide; ACQ-6=Asthma Control Questionnaire; AQLQ=Asthma Quality-of-Life Questionnaire.

Figure 1 (top panel) shows box and whisker plots with males (blue) and females (red) identified the mean RBC:membrane significant difference (*P*=.01) in patients with moderate-severe asthma. The middle panel shows significant sex-dependent linear relationships for RBC:membrane with age in healthy (ρ=-.55, *P*=.002, Y=-0.0035x+0.5776) and asthma (ρ=-.49, *P*=.006, Y=-0.0022x+0.4484) participants. A significant ANCOVA model revealed the significant influence of age (*P*=.005), sex (*P*<.001), hemoglobin (*P*=.008), and asthma status (*P*=.02) on RBC:membrane values. As shown in Figure 2, significant relationships for RBC:membrane with hemoglobin (ρ=.29, *P*=.04) and DL_CO_ (ρ=.71, *P*<.001) were also observed.

**Figure 1.**
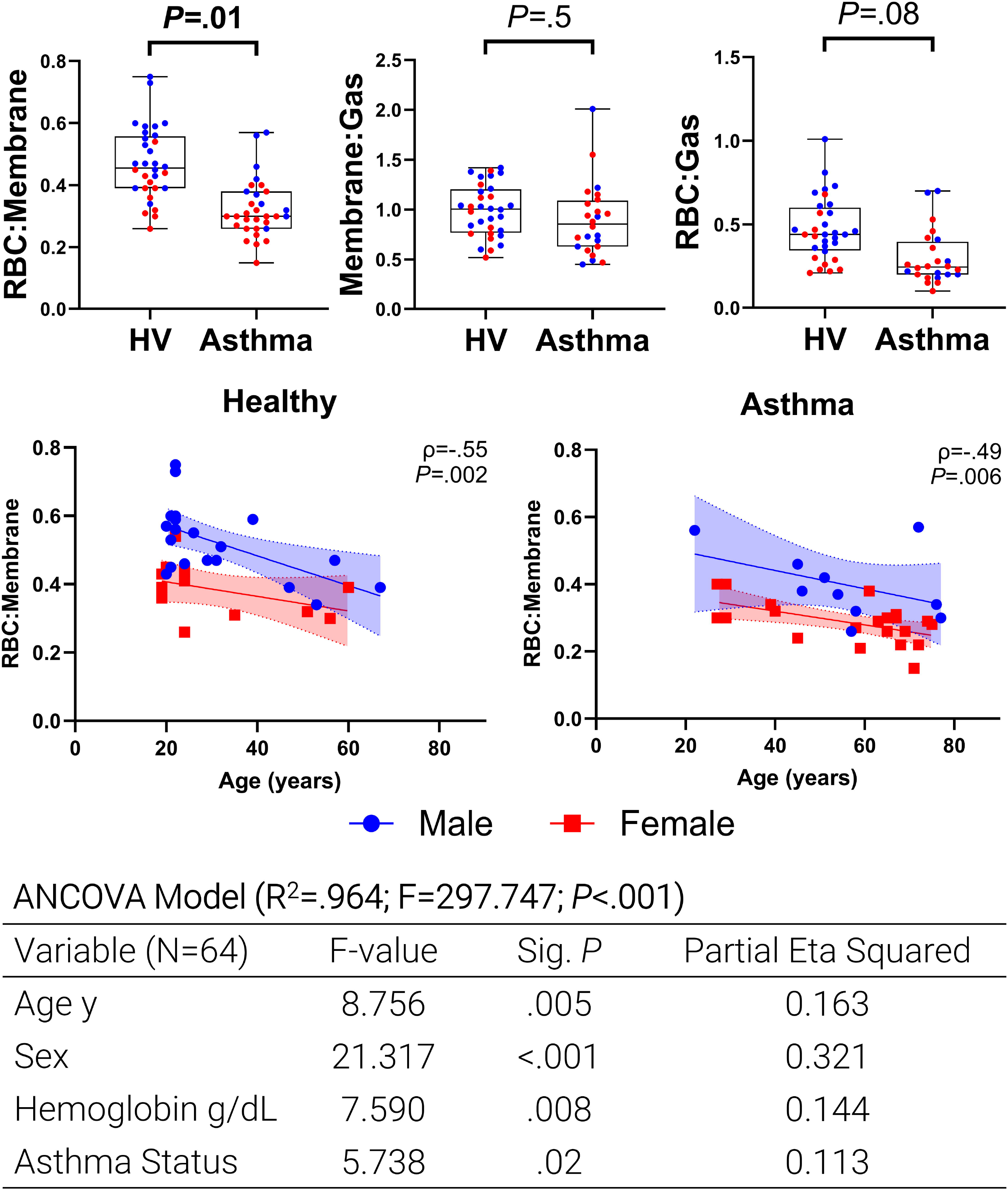
^129^Xe MRI RBC:membrane in Asthma and relationships with age & sex. Box and whisker plots show there was significantly different RBC:Membrane (*P*=.01), but not Membrane:Gas (*P*=.5) or RBC:Gas (*P*=.08) in healthy and asthma participants. Box represents mean, whiskers represent min–max range. Significant relationship with age for RBC:membrane for healthy (ρ=-.55, *P*=.002, Y=-0.0035x+0.5776) and asthma participants for RBC:Membrane (ρ=-.49, *P*=.006, Y=-0.0022x+0.4484). *P*=Significance values for ANCOVA adjusted for sex. ANCOVA model shows significant effects on RBC:membrane of age (P=.005), sex (P<.001), hemoglobin (P=.008), and asthma status (P=.02). The model’s explanatory power is high (R²=.964; F=297.747; P<.001). *BD=bronchodilator; RBC=red blood cell*.

**Figure 2.**
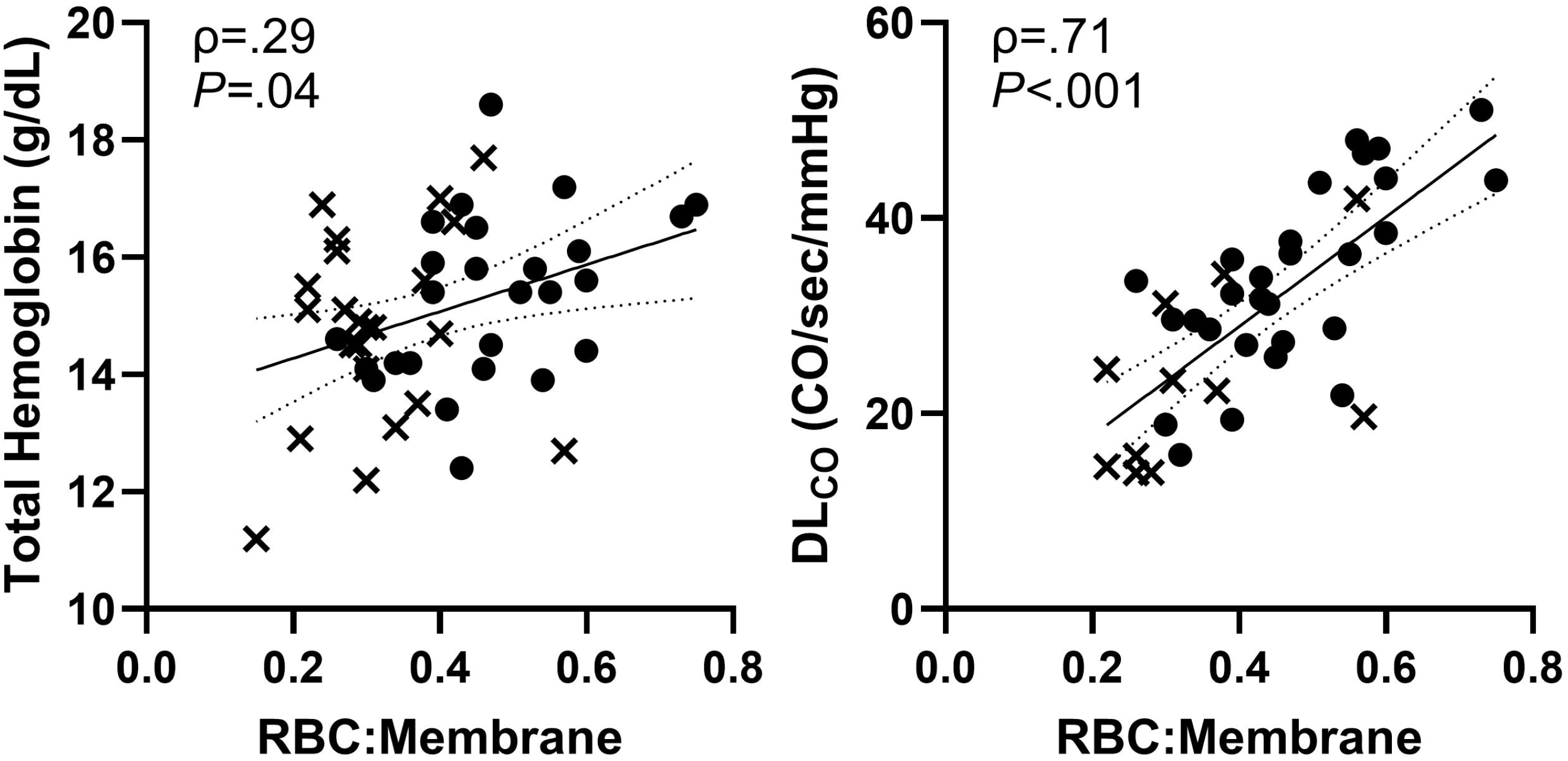
RBC:membrane Relationships. Significant relationships for RBC:membrane with pulse-oximetry measured hemoglobin (ρ=.29, *P*=.04, y=4.00x+13.48) and DL_CO_ (ρ=.71, *P*<.001, y=56.06x+6.50). *P*=Significance values. *RBC=red blood cell*; *DL*_*CO*_*=diffusing capacity of the lungs for carbon monoxide*.

## DISCUSSION

^129^Xe MRI offers novel insights into airway, terminal airway and gas-transfer function in patients with asthma because of its modest Ostwald solubility in biologic membranes, including the alveolar and RBC membranes. Once inhaled, ^129^Xe gas rapidly flows through airways and to terminal airways throughout the lung to provide reproducible measurements of airways dysfunction. ^129^Xe also participates in alveolar transmembrane diffusion and binds to microvascular RBC, displacing molecular oxygen on hemoglobin-bound heme. Because the ^129^Xe gas, alveolar membrane and RBC signals resonate at different frequencies, their individual resonance peaks may be quantified directly using the area-under-the-curve (AUC) and normalized to one another. The ratio of the RBC signal peak AUC to the alveolar membrane signal peak AUC results in RBC:membrane, which has been proposed as a voxel-specific surrogate of gas-transfer.

We recently discovered that bulk blood volume in the large- (BV_10_) and small-vessels (BV_5_)^4^ differed in severe asthma, compared to healthy volunteers, which was consistent with the notion of pulmonary vascular pruning in severe asthma,^4^ hypoxic vasoconstriction and/or vascular wall structural remodeling.^2^ It is impossible to undertake serial histology to determine the temporal dynamics of these processes, but ^129^Xe MRI makes this possible non-invasively, *in vivo*, with high spatial-temporal resolution, without radiation, and with obvious advantages over histology. To better understand the potential influence of pulmonary vascular abnormalities on gas-exchange in patients with moderate-severe asthma, here we evaluated a small group of patients and healthy volunteers using ^129^Xe MR RBC:membrane and pulmonary function tests. We observed: 1) mean ^129^Xe RBC:membrane was significantly different in asthma compared to the healthy volunteers (*P*=.01), 2) RBC:membrane values were both sex- and age-dependent in healthy (*P*=.002) and asthma participants (*P*=.006) participants, and, 3) in a significant model, age (*P*=.005), sex (*P*<.001), hemoglobin (*P*=.008), and asthma status (*P*=.02) influenced RBC:membrane values.

The difference between asthma and healthy volunteer RBC:membrane values, even after age-correction, was unexpected. The fact that Membrane:gas did not differ but there was a trend towards diminished RBC:gas values in asthma participants, suggests RBC pulmonary vascular differences may be responsible for the abnormal RBC:membrane values in asthma. This hypothesis is supported by the finding of abnormally diminished small vessel blood distribution (coined pruning) observed in the Severe Asthma Research Program.^4,5^ Lending support to the finding of lower RBC:membrane in asthma participants, a significant ANCOVA model also revealed the contributions of sex, age, hemoglobin and asthma to RBC:membrane.

Similar to previous work, we observed an age-related decrease in the RBC:Membrane ratio.^8-10^ In addition, the finding of moderate relationships for all participants between RBC:membrane and both DL_CO_ and hemoglobin is consistent with our understanding of gas-exchange and the role of hemoglobin and heme binding by ^129^Xe atoms.

In conclusion, we report significantly different ^129^Xe RBC:membrane values in moderate-severe asthma participants as compared to healthy volunteers, which may be related to pulmonary vascular remodeling in these patients.

## Data Availability

All data produced in the present work are contained in the manuscript.

